# Epidemiological and molecular evidence of intrafamilial transmission through sexual and vertical routes in Bahia, the state with the highest prevalence of HTLV-1 in Brazil

**DOI:** 10.1101/2022.12.06.22283194

**Authors:** Aidê Nunes da Silva, Thessika Hialla Almeida Araújo, Ney Boa-Sorte, Giovanne Farias, Ana Karina Galvão-Barroso, Antônio de Carvalho, Ana Carolina Vicente, Bernardo Galvão-Castro, Maria Fernanda Rios Grassi

## Abstract

**Introduction:** Familial clustering of HTLV-1 and related diseases has been reported in Brazil. However, intrafamilial transmission of HTLV-1 based on molecular analysis has been studied only in few communities of Japanese immigrants and African-Brazilians.

**Objective:** To investigate the familial clustering of HTLV-1 infection and to determine the likely routes of transmission through epidemiological and genetic analyzes.

**Methods:** Medical records of 1,759 HTLV-1+ patients from de the Center for HTLV in Salvador, Brazil, were evaluated to identify first-degree relatives previously tested for HTLV-1. Familial clustering was assumed if more than one member of the same family was HTLV-1+. LTR regions of HTLV-1 sequences were analyzed for the presence of intrafamilial polymorphisms. Family pedigrees were constructed and analyzed to infer the likely transmission routes of HTLV-1.

**Results:** In 154 patients at least one other family member had tested positive for HTLV-1 (a total of 182 first-degree relatives). Of the 91 couples (182 individuals), 51.6% were breastfed, and 67.4% reported never using a condom. Of the 42 mother-child pairs, 23.8% had a child aged 13 years or younger; all mothers reported breastfeeding their babies. Pedigrees of families with 4 or more members suggests that vertical transmission is a likely mode of transmission in three families. Three families may have had both vertical and sexual transmission routes for HTLV-1. The genetic signatures of the LTR region of 8 families revealed 3 families with evidence of vertical transmission, another 3 families (spouses) with sexual transmission, and one family with both transmission routes. HTLV-1 sequences belonged to Cosmopolitan subtype HTLV-1a Transcontinental subgroup A. Conclusion: Sexual and vertical transmission routes contribute to the intrafamilial spread of HTLV-1 in the state of Bahia.

**AUTHOR SUMMARY:** Human T-lymphotropic virus type 1 (HTLV-1) was the first human retrovirus isolated in the early 1980s. It is estimated that approximately 10 million people worldwide are currently infected with HTLV-1, and most people living with HTLV (PLwHTLV) live in developing countries. The virus is associated with a wide range of diseases, including neoplasms such as adult T-cell leukemia/lymphoma and progressive and disabling myelopathy, but most PLwHTLV are unaware of their serologic status. HTLV-1 is transmitted through contact with contaminated blood and derivatives, sexually, and from mother to child, especially through breastfeeding. Only recently has WHO recognized HTLV-1 as a as threatening pathogen to human, but in many parts of the world HTLV screening is not performed in blood banks or in pregnant women. This may promote silent intrafamilial transmission of the virus across generations and promote familial clustering of the virus and associated diseases. In this study, we investigated the familial clustering of HTLV-1 infection in the state of Bahia, an endemic area for this virus in Brazil. We found that both sexual and vertical pathways contribute to the transmission and persistence of the virus in families across multiple generations. Therefore, in addition to expanding screening for pregnant women and providing infant formula to infected mothers, it is of utmost importance to combat sexual transmission through effective measures that can help address this serious and neglected public health problem.

## INTRODUCTION

Human T-lymphotropic virus type 1 was the first human retrovirus to be described (1). HTLV-2, which is rarely associated with disease, was then isolated soon after (2). HTLV-1 is the causative agent of adult T-cell leukemia/lymphoma (ATLL) (3), tropical spastic paraparesis/ HTLV-1-associated myelopathy (TSP/HAM) (4,5), HTLV-1-associated uveitis (HAU) (6) and HTLV-1-associated infectious dermatitis (HAID) (7). In addition, many other diseases have been associated with HTLV-1 infection, including polymyositis, sinusitis, bronchoalveolar pneumonia, keratoconjunctivitis sicca and bronchiectasis, suggesting multisystemic involvement (8–10).

The worldwide prevalence of HTLV-1 infection is estimated to be between 5 and 10 million cases (8). Of all affected countries, Brazil may be the one with the highest absolute number of persons living with HTLV-1 (PLwHTLV) (8,11). Salvador, the capital of the state of Bahia, located in northeastern Brazil, has the highest prevalence in the country (12). A representative sampling of this city’s general population revealed a seroprevalence of 1.8%, corresponding to around 50,000 HTLV-infected individuals (13). An ecological prevalence study published in 2019 reported that HTLV-1 is widespread throughout the state of Bahia, and that at least 130,000 individuals were infected with the virus (14).

Horizontal transmission through sexual contact appears to be the main route of HTLV-1 infection in the general population of Salvador (15). However, there is evidence that vertical transmission also occurs in the state of Bahia, as the prevalence of HTLV-1 infection in pregnant women has been consistently estimated at around 1%(16–18); moreover, approximately 2% of HTLV-1-infected individuals are reportedly aged 15 years or younger (14).

The prevalence of HTLV-1 is heterogenous throughout the country, with higher prevalence noted in some geographic regions, especially northeastern Brazil (19). HTLV-1 infection tends to be more predominant with age, with females being more affected (8). The familial aggregation of HTLV-1 infection, as well as its associated diseases, has been reported worldwide, including in Brazil (18,20–23). However, in this country, the intrafamilial transmission of HTLV-1 based on molecular analysis has only been studied in communities of Japanese immigrants and their descendants (24–26) (and in isolated communities of African descent (27). This study aims to investigate familial clustering of HTLV-1 infection in the state of Bahia, as well as to determine likely routes of transmission through epidemiological and genetic analysis.

## METHODS

### Study design and population

The present cross-sectional study was conducted between February 2017 and November 2021 at the Integrative and Multidisciplinary Center for HTLV (CHTLV), Bahia School of Medicine and Public Health Salvador (EBMSP), in Salvador, Bahia-Brazil. This outpatient clinic provides comprehensive biopsychosocial care to the public and is supported by the public health services of the Brazilian National Health System (Sistema Único de Saúde [SUS]) (28).

The medical records of 1,759 individuals were evaluated to identify first-degree relatives (parents, children, siblings and spouses) who had previously been tested for HTLV-1 by ELISA, with confirmation by Western blotting (Figure 1). Families with at least two members (including the patient) who had been tested were included in the study. Patients and family members with cognitive impairment, psychiatric/psychological disorders, and those diagnosed with HTLV-2 and/or coinfected with HIV, HBV or HCV were excluded. Familial aggregation was considered when more than one member of the same family tested positive for HTLV-1, with the index case being the first family member who was registered at CHTLV.

**Fig 1.**
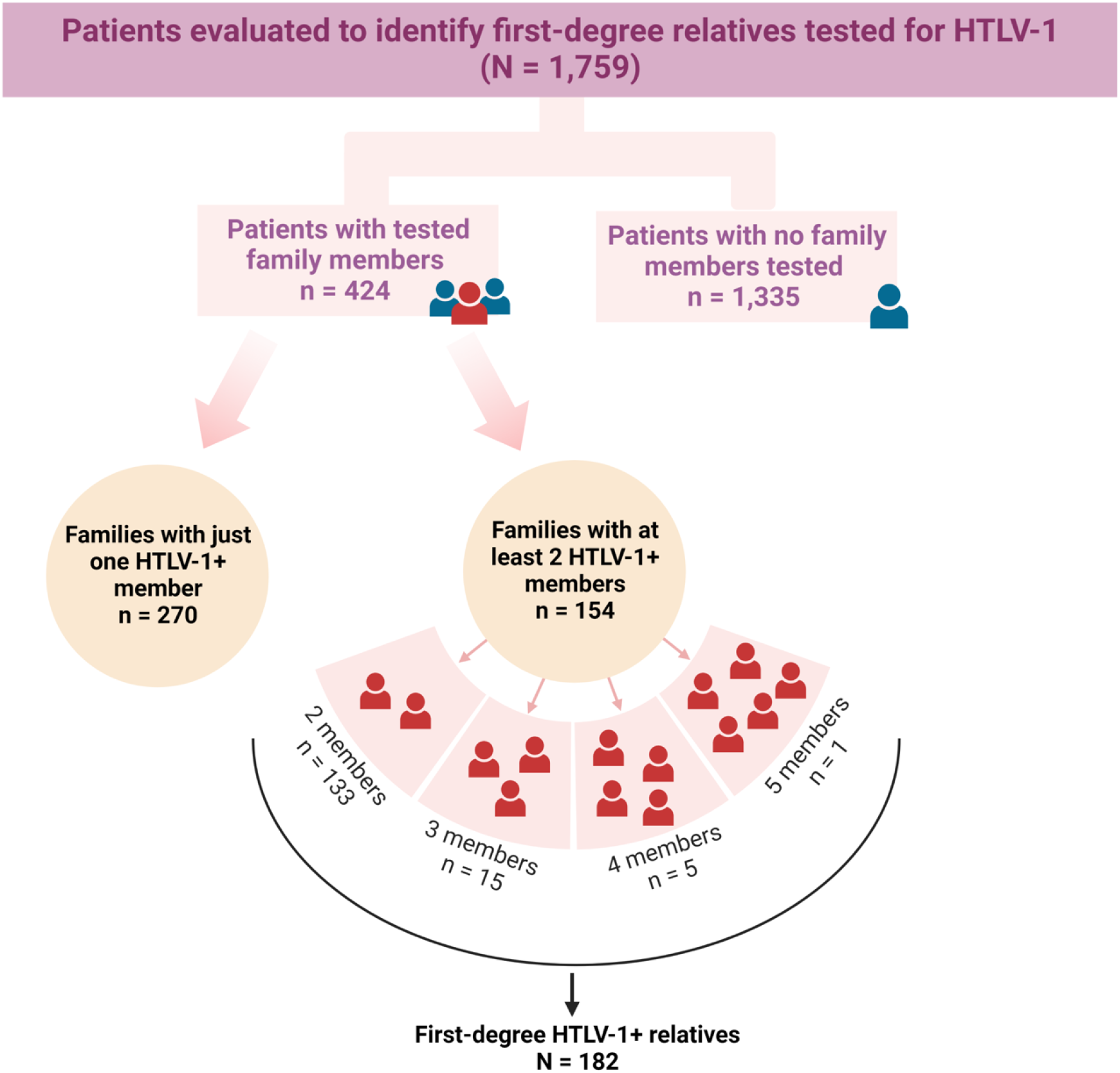
Study design flowchart.

Semi-structured questionnaires were used to collect sociodemographic, clinical, and behavioral data. Patients with TSP/HAM were diagnosed according to previously described criteria (29).

### Genetic sequencing

Blood samples from eight index cases and 12 HTLV-1 seropositive family members were collected for DNA extraction using a QIAamp DNA Blood Mini Kit (Qiagen, Hilden, Germany) in accordance with the manufacturer’s instructions. The HTLV-1 long terminal repeat (LTR) region was amplified by nested PCR using a previously described protocol (30). PCR products were purified using a QIAamp PCR purification kit (Qiagen). Direct nucleotide sequencing was performed in both directions using a Big Dye Terminator v 3.1 Cycle Sequencing Ready Reaction Kit (Applied Biosystems, Foster City, CA) on a 3100 Automated DNA Sequencer (Applied Biosystems). Subsequently, the LTR sequences were uploaded to the HTLV-1 and 2 Genotyping Tool to identify HTLV-1 subtype (31). All HTLV-1 LTR sequences were edited and aligned using BioEdit v5.0.9 software (Department of Microbiology, North Carolina State University, USA) and then visually analyzed to detect the presence of intrafamilial polymorphisms. The new nucleotide sequences identified in the present study were deposited in the GenBank database under accession numbers OP831963-OP831989.

### Data analysis

Descriptive statistics are reported as means ± SD for continuous variables, or frequencies (%) for categorical variables. The student’s t test and chi-square test or Fisher’s exact test were used for comparisons. The family pedigrees were generated and analyzed to infer the likely routes of HTLV-1 transmission between family members. All data were analyzed using SPSS v20 software (SPSS Inc, Chicago, USA). Statistical significance was assumed when p < 0.05.

## RESULTS

We identified 424 HTLV-1+ patients with at least one first-degree relative previously serologically tested for HTLV-1; of these, 154 patients (index cases) had at least one other family member who was positive for HTLV-1. In all, 182 first-degree relatives of index cases tested positive for HTLV-1 antibodies (Figure 1). The prevalence of HTLV-1 in family members was estimated at 32.9% (182/553 tested family members). The overall prevalence of familial clustering of HTLV-1 was 36.3% (154/424; CI95%:31.7-41.1). The mean ages (SD) of the index cases and their family members were 44.2 (16.12) years and 41.1 (18.64) years, respectively (p =0.781). Females were more prevalent among both index cases and family members (p=0.009). Most individuals self-reported brown or black skin color. Many family members, 40.9% (63/154) of index cases and 39% (71/182) of first-degree relatives, had less than seven years of formal schooling (Table 1). The prevalence of TSP/HAM was higher in index cases (61/154; 39.6%) compared to infected family members (37/182; 20.3%) (p < 0.001).

**Table 1.** Composition of family units of HTLV-1-infected patients (index cases).

| Family composition | No. of families | No. of individuals |
| --- | --- | --- |
| **Two-member families** | **133** | **266** |
| IC + spouse | 91 | 182 |
| Mother + child | 42 | 84 |
| **Three-member families** | **15** | **45** |
| IC + current wife + ex-wife | 2 | 6 |
| IC + husband + son/daughter | 7 | 21 |
| IC + sons | 2 | 6 |
| IC + husband + mother | 1 | 3 |
| IC + mother + father | 1 | 3 |
| IC + mother + son | 1 | 3 |
| **Four-member families** | **5** | **20** |
| IC + husband + 2 children | 1 | 4 |
| IC + 3 children | 2 | 8 |
| IC + wife + 2 children | 1 | 4 |
| IC + mother + 2 children | 1 | 4 |
| **Five-member families** | 1 | 5 |
| IC + mother + father + son + daughter | 1 | 5 |
| **Total** | **154** | **336** |

IC: Index case

In 133/154 families with HTLV-1 aggregation, two members were found to be infected: 91 (68.4%) were husband and wife, while 42 (31.6%) were mother and child. In addition, there were three infected persons in 15 families, while five families had four HTLV-1+ members, and in one family five individuals tested positive (Figure 1, Table 2).

**Table 2.** Sociodemographic and clinical profile of index cases and family members with HTLV-1.

| Variable | Index case | Family members | p value* |
| --- | --- | --- | --- |
|  | N (%) | N (%) |  |
|  | 154 | 182 |  |
| Sex |  |  | 0.009 |
| Male | 47 (30.5) | 81(44.5) |  |
| Female | 107(69.5) | 101(55.5) |  |
| Age (years) |  |  | 0.059 |
| 0 to 19 | 9 (5.84) | 26 (14.3) |  |
| 20 to 39 | 51 (33.11) | 58 (31.8) |  |
| 40 to 59 | 71 (46.1) | 68 (37.3) |  |
| 60 to 79 | 20 (12.9) | 28 (15.3) |  |
| 79+ | 3 (2.0) | 2 (1.09) |  |
| Skin color (self-reported) |  |  | 0.542 |
| Black or mixed-race | 107 (69.4) | 121 (66.5) |  |
| White | 12 (7.8) | 11 (6.0) |  |
| Missing information | 35 (22.8) | 50 (27.5) |  |
| Marital status |  |  | 0.331 |
| Single | 36 (23.3) | 47 (26.7) |  |
| Married/Stable relationship | 106 (68.8) | 110 (60.4) |  |
| Divorced | 4 (2.6) | 5 (2.74) |  |
| Widowed | 6 (3.89) | 13 (7.14) |  |
| Missing information | 2 (1.29) | 7 (3.84) |  |
| Years of education |  |  | 0.934 |
| ≤ 7 Years | 63 (40.9) | 71 (39.0) |  |
| > 7 Years | 85 (55.2) | 94 (51.7) |  |
| Missing information | 6 (3.9) | 17 (9.3) |  |
| TSP/HAM diagnosis | 61 (39.6) | 37 (20.3) | 0.001 |

\*Chi-squared test

Of the 91 HTLV-1+ couples (spouses), totaling 182 individuals, 51.6% (94/182) reported being breastfed, and the majority (67.4%; 122/182) reported never using a condom. Of the 42 mother-child pairs, 10 (23.8%) children were aged 13 years or younger, all mothers reported breastfeeding their babies, and none of the mothers, nor their children, had received blood transfusions. The evaluation of pedigrees of families with three or more HTLV-1+ members suggests the likelihood of vertical transmission in seven families (#22, #30, #36, #77, #87 #101, #124) (Figure 2), sexual transmission in two families (#2, #4) (Figure 2), while both vertical and sexual routes of HTLV-1 transmission may have occurred in eleven families (#9, #32, #65, #80, #97, #114, #120, #140, #142, #144, #146) (Figure 2). In eight families, genetic sequencing of the LTR region revealed strong evidence of vertical transmission in three families (#153, #154 and #158), possible horizontal (sexual, wife and husband) transmission in another three families (#155, #156 and #157), and one family with evidence of both types of transmission (family #159). In one family (#152), no unique genetic signature was identified (Figure 3). In all family members, sequencing of the HTLV-1 LTR region identified HTLV-1 Cosmopolitan subtype Transcontinental subgroup A.

**Fig 2.**
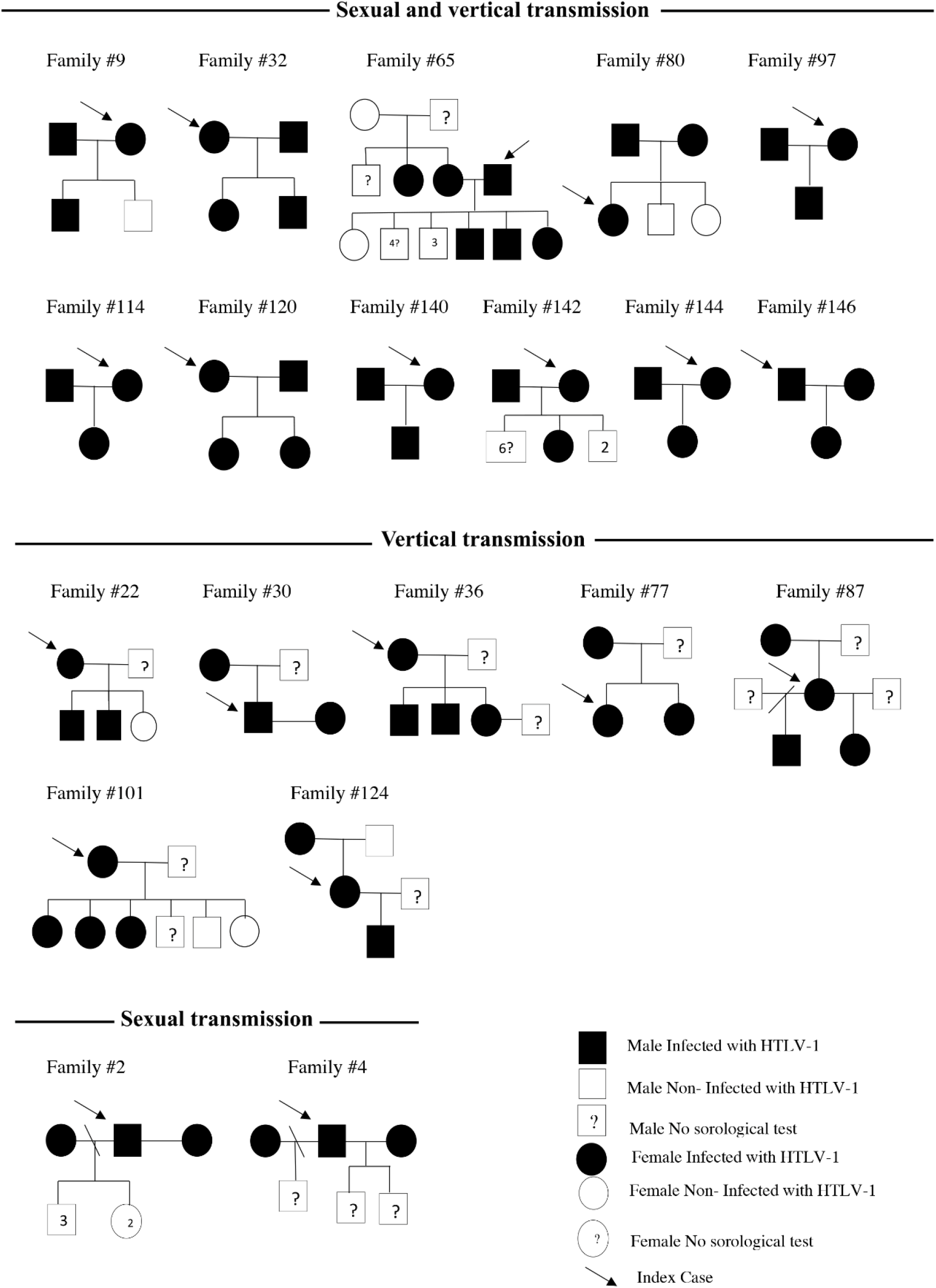
Pedigree of 20 family nuclei and possible transmission routes of HTLV-1 based in epidemiological data.

**Fig 3.**
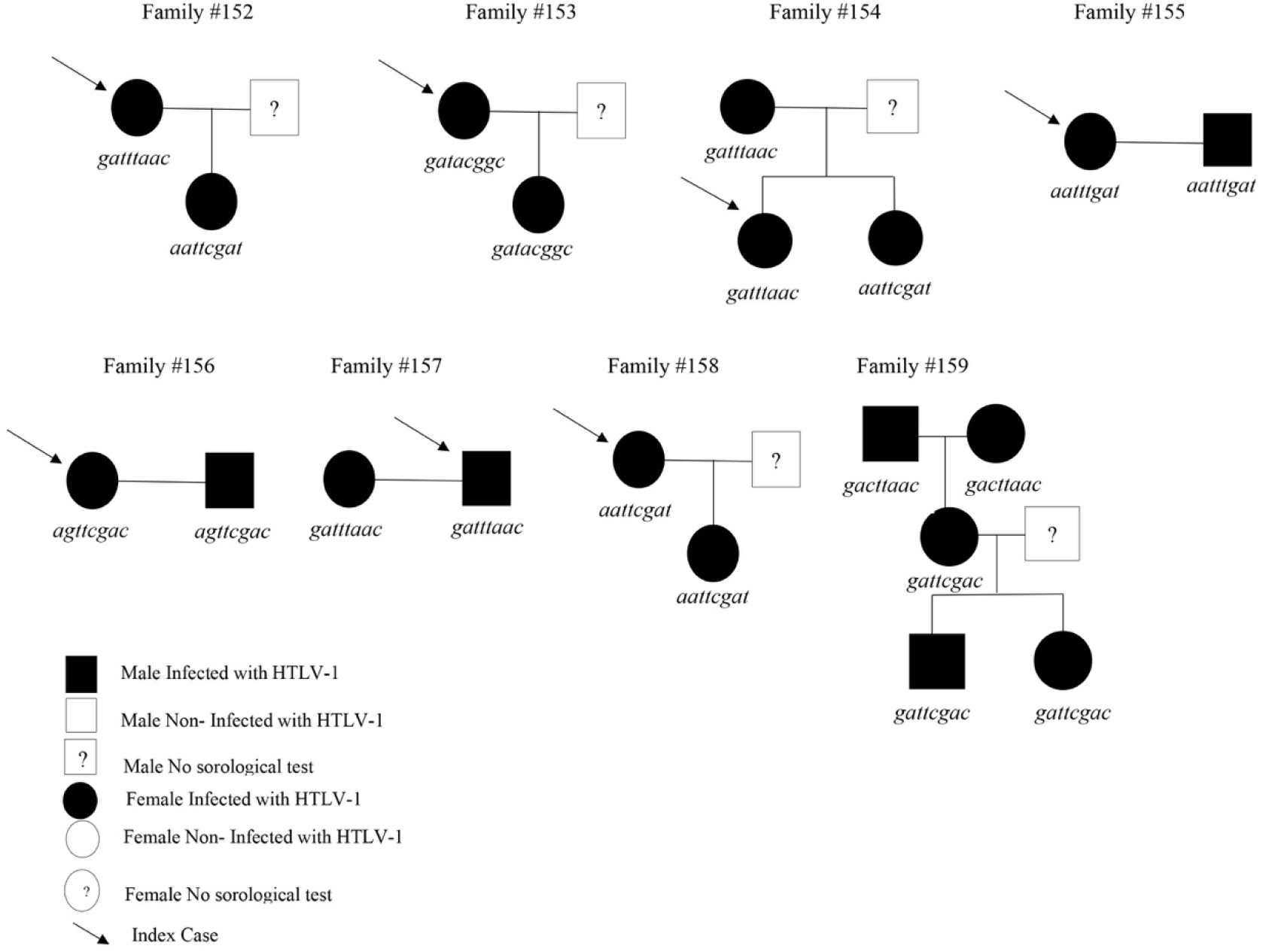
Pedigree of 8 family nuclei and possible transmission routes of HTLV-1 based in genetic sequencing of the LTR region.

## DISCUSSION

The present study employed epidemiological and molecular analysis in an attempt to elucidate the sexual and vertical pathways underlying the intrafamilial spread of HTLV-1 in the state of Bahia. Importantly, both vertical and horizontal transmission were concomitantly observed in some families.

The overall prevalence of HTLV-1 familial aggregation found herein was 36.3% CI95%:31.7-41.1), a rate similar to that observed in other endemic areas in countries such as Taiwan (38.8%) and Argentina (31.5%) (32,33), yet higher than that observed in Benin, West Africa (27.5%) (34).

Considering the prevalence of HTLV-1 familial aggregation throughout Brazil, the present rate (36.3%) is slightly lower than that previously reported in patients studied at an HTLV-1 center in the country’s northern region (43.5%) (23). A previous study conducted in Bahia examined 43 family members of 20 HTLV-1-infected pregnant women, and reported a prevalence of 32.6%, suggestive of familial clustering (18). In consonance, the prevalence of HTLV-1 among family members tested herein was estimated at 32.9%. The populations served by HTLV centers possess similar sociodemographic profiles, e.g., a predominance of women with low income and educational levels compared to blood donors (35). In contrast, a study investigating the family members of index cases recruited from blood donors in the southern region of Brazil identified a slightly lower prevalence (25.9%) (22).

Regarding the pathways of intrafamilial spread of HTLV-1 in Brazil, studies conducted in communities with a high prevalence of HTLV-1, such as those with Japanese migrants and their descendants (25,26) or Brazilians of African descent living in isolated communities (27), suggested that both vertical and sexual transmission contribute to the maintenance of HTLV-1 spread across generations. The present study also found evidence of both modes of transmission, even occurring simultaneously in some of the families investigated (#32, #65, #159).

As vertical transmission is considered the main route of intrafamilial spread of HTLV, preventive measures have been taken to address this concern. Measures implemented in Japan to control vertical HTLV-1 transmission have successfully decreased prevalence significantly (36). In Bahia, HTLV-1 testing for pregnant women became mandatory in 2011 (Portaria No 460, de 19 de novembro de 2020) and has more recently been recommend across Brazil (37). The World Health Organization and Pan American Health Organization have both strongly recommended mandatory testing for pregnant women in recognition of the importance of vertical transmission (38). However, little attention has been paid to the intrafamilial spread of HTLV-1 through sexual intercourse (39). Recently, an increase in the rate of transmission of HTLV-1 through sexual intercourse, particularly among young people was observed in Japan (40). Studies conducted in Salvador have identified sexual transmission as the main route responsible for HTLV-1 in the general population (13,15); moreover, approximately 1% of pregnant women tested in Salvador were found to be infected with HTLV-1 (16,17).

In addition, the familial aggregation of HTLV-1 has also been associated with the clustering of diseases related to the virus. A systematic review investigating HTLV-1-associated diseases reported a higher incidence of ATLL and TSP/HAM in relatives of index cases (20). The present study also identified TSP/HAM in approximately 20% of the relatives of HTLV-1+ patients. Moreover, previous studies conducted in Salvador have additionally identified familial clusters of IDH in association with TSP/HAM (21,41). Another study further demonstrated the familial aggregation of TSP/HAM, with onset occurring a younger age (41).

The HTLV-1 subtype identified in the studied sample belongs to the Cosmopolitan subtype Transcontinental subgroup A (HTLV-1a). This subtype was also detected in previous studies of populations in Bahia, supporting the introduction of the virus during the post-Columbian period through the slavery trade (43,44).Conversely, Japanese subgroup (a) was reported in a subset of samples from Japanese immigrants and their families, confirming two different pathways of HTLV-1 introduction in Brazil during the post-Colombian period (25,26).

Our study presents some limitations with respect to bias in sample selection, as the sample was mainly composed of adults with an average age of 50 years. In addition, the percentage of tested family members was low, with approximately 1.2 relatives tested per index case, and spouses representing the majority of family units in which familiar aggregation was observed.

In conclusion, the results presented herein demonstrate that both sexual and vertical transmission routes contribute to the intrafamilial spread of HTLV-1 in the state of Bahia.

Therefore, we strongly recommend testing all family members of HTLV-1-infected individuals to better understand relevant factors related to the interfamilial spread of this virus. Considering that both vertical and sexual routes play a role in the spread of HTLV-1 (33), in addition to expanding the screening of pregnant women and providing infant formula to infected mothers, it is also of paramount importance to address sexual transmission by implementing effective measures, which may contribute to the control this serious and neglected public health problem that mainly affects socio-economically disadvantaged populations.

## Data Availability

The raw data supporting the conclusions of this article will be made available by the authors, without undue reservation.

## ETHICS STATEMENT

The studies involving human participants were reviewed and approved by the Comitê de Ética em Pesquisa em Seres Humanos da Bahiana (CAAE: 60554416.8.0000.5544). Written informed consent from the patients/participants or their legal guardian/next of kin was not required to participate in this study in accordance with the national legislation and the institutional requirements.

## AUTHOR CONTRIBUTIONS

BG-C and MFRG: conceptualization. MFRG ANS, THAA, GF, AK G-B, AC, ACV, and NB-S: data curation. ANS, THAA, NBS ACV, BG-C and MFRG: formal analysis and methodology. All authors involved in investigation, writing—original draft, review and editing, contributed to the article, and approved the submitted version.

## ACKNOWLEDGMENT

We would like to thank Sônia Rangel and Maíara Cerqueira for administrative support and Dr Fred Neves Santos for critically reading the manuscript. We would also like to thank Andris K. Walter for critical analysis, English language revision, and manuscript copyediting assistance.

